# Ketamine Modulates the Neural Correlates of Reward Processing in Unmedicated Patients in Remission from Depression

**DOI:** 10.1101/2020.12.07.20230151

**Authors:** Vasileia Kotoula, Argyris Stringaris, Nuria Mackes, Ndabezinhle Mazibuko, Peter.C.T Hawkins, Maura Furey, H Valerie Curran, Mitul.A. Mehta

## Abstract

Ketamine as an antidepressant improves anhedonia, a pernicious symptom of depression as early as 2h post-infusion. The effects of ketamine on anhedonia are thought to be exerted via actions on reward-related brain areas—yet, these actions remain largely unknown. This study examines ketamine’s effects during the anticipation and receipt of an expected reward, after the psychotomimetic effects of ketamine have passed, when early antidepressant effects are reported. In order to identify brain areas that are modulated by the drug per se and are not linked to symptom changes, we have recruited 37 participants who remitted from depression and were free from symptoms and antidepressant treatments at the time of the scan. Participants were scanned while performing a monetary reward task and we examined ketamine’s effects on pre-defined brain areas that are part of the reward circuit. An overall effect of ketamine was observed during the anticipation and feedback phases of win and no-win trials. The drug effects were particularly prominent in the nucleus accumbens and putamen, upon the receipt of smaller rewards and the levels of (2R,6R)-HNK, 2h post-infusion, significantly correlated with the activation observed in the ventral tegmental area (VTA) for that contrast. These findings demonstrate that ketamine can produce detectable changes in reward-related brain areas, 2h after infusion, which occur without symptom changes and support the idea that ketamine might improve reward-related symptoms via modulation of response to feedback.

## Introduction

MDD is characterised by altered reward processing and a reduced ability to modulate behaviour as a function of rewards (1). Deficits in reward processing can precede the onset of depression(2), are linked to anhedonia and persist during remission (3, 4). Ketamine, an NMDA receptor antagonist, produces robust antidepressant effects that occur as early as 2h after drug infusion, peak at 24h and last up to one week (5). In relation to reward processing, the drug improves anhedonia, a symptom known to be resistant to standard anti-depressant treatment(6). To our knowledge however, no study has examined whether ketamine’s ability to improve anhedonia is the result of direct modulation of reward processing areas that is not secondary to changes in symptoms. In this study, we have used a well-validated fMRI task, the monetary incentive delay (MID) task (7) in order to examine whether the drug engages brain areas involved in reward processing, two hours after its administration, in a large sample of treatment-free and symptom-free remitted depressed volunteers.

In the brain, reward processing is mainly subserved by regions that are part of the mesocorticolimbic pathway (8). Imaging studies that have used the MID task to examine reward processing in healthy volunteers showed that striatal regions, especially the caudate and the putamen, but also the insula and frontal brain areas are activated during the anticipation phase of the MID, when a monetary reward is expected (9). During the feedback phase of the task when the expected reward is delivered, a similar set of brain regions appear to be involved (10). In depression, recent meta-analyses showed that the ventral striatum (VS), the caudate and the putamen present with decreased activation during the anticipation and feedback phases of the MID (2, 11-14). This hypoactivation of reward processing areas observed in depression also persists in remission with studies indicating that compared to healthy controls remitted depressed volunteers show blunted responses to reward (3) and decreased activation in prefrontal (4) and striatal (15) regions during loss anticipation and outcomes. Given the central role of reward processing in depression, compounds that target these areas are considered promising candidates for alleviating depression, including anhedonia(16).

Ketamine improves anhedonia as early as two hours after a single infusion, although the neural basis of these effects is only beginning to be understood. Using [18F]FDG-PET imaging at two hours post dosing, glucose metabolism in the dorsal anterior cingulate cortex (dACC) and the putamen correlated with reduced anhedonia in patients with treatment-resistant bipolar depression (17). In MDD patients, reductions in anhedonia correlated with increased glucose metabolism in the dACC and hippocampus (18). Anhedonia is not a unitary construct with separable components including reward anticipation and feedback or delivery (19) as measured by the MID. Research in non-human primates suggests that ketamine treatment could ameliorate blunted anticipatory responses to appetitive stimuli by normalizing brain activation in the sub-genual anterior cingulate cortex (sgACC) (20). One study in patients with depression investigated ketamine-induced changes in brain activity and anhedonia using a reward-related fMRI task, demonstrating a reduction in sgACC hyperactivity to positive feedback in 14 patients tested within five days of a ketamine infusion (21). The fact that the changes in the metabolism and activation of reward associated brain areas temporally overlap with symptom changes makes it difficult to determine whether these changes are due to the primary effects of the drug or are secondary to the effect of ketamine on depressive symptoms. While these PET and fMRI studies provide insights about the neural mechanisms that accompany ketamine’s early antidepressant action, the effects on brain regions associated with anticipatory and feedback components of reward tasks during the emergent period of the antidepressant response (2-24hours) are not known.

At a neuronal level, ketamine and its main metabolite, norketamine, indirectly activate the post-synaptic AMPA receptors and trigger molecular pathways, including BDNF and mTOR pathways that lead to an increase in synaptic plasticity, which has been linked to the antidepressant effects of the drug. (for review see (22)). Another metabolite, (2R,6R)-HNK can bind and activate AMPA receptors directly, and thus trigger the initiation of plasticity-related molecular processes (23). In animal models of anhedonia, changes in plasticity markers following ketamine have been linked to increased activations of the reward pathways that are mainly mediated by dopamine (for review see (24)). While direct actions of (2R,6R)-HNK are a candidate for such improvements, its action as an antidepressant remains to be tested in humans and links between this metabolite and anhedonia related changes in brain activations have yet to be observed.

In this study, we aimed to investigate the effects of ketamine on task performance and functional brain response to the MID task two hours post-infusion – the time at which early antidepressant effects are reported – in a cohort of participants who remitted from depression -. We chose to recruit treatment free, remitted depressed participants since they present with altered brain activations in reward related areas (3, 25, 26) that might resemble those observed in depression and would allow the examination of ketamine’s effects without the confounds of antidepressant treatment or concurrent symptom change. We have focused on specific regions of interest associated with reward processing who are activated during the MID task, namely the striatum, the VTA, the amygdala and the insula (2, 9-12). We hypothesise that ketamine would increase activation in those areas. We also examine cortical areas associated with reward in a exploratory whole brain analysis. Moreover, we measured the levels of ketamine’s metabolites to explore whether norketamine and (2R, 6R)-HNK levels correlate with any ketamine related changes in the activation of reward processing brain areas.

## Methods

### Participants

37 remitted depressed volunteers (21 female, mean age= 28.5 years) took part in a randomised double-blind, placebo controlled, cross-over study. The MINI International Neuropsychiatric Interview was used to confirm history of depression and remission at study entry. Inclusion criteria included a minimum of three months of no antidepressant treatment, prior to taking part. The exclusion criteria included any history of other psychiatric or neurological disorder, a previous adverse response to ketamine; any medical conditions that affect hepatic, renal or gastrointestinal functions; cardiac abnormalities; hypertension; a significant history of substance abuse or a positive test for drugs of abuse at screening or a study day; nicotine use (≥5 cigarettes per day), alcohol (≥28 units/week) and caffeine (≥ 6 cups per day) or any MRI contraindications. All participants gave written informed consent for the study, which was approved by the Psychiatry, Nursing and Midwifery Research Ethics Subcomittee (reference: HR-14/15-0650)

### Study procedures

Participants who met eligibility criteria were randomized to receive either a single intravenous infusion of ketamine (0.5 mg/kg) or placebo (0.9% saline solution) during the first session and the alternative treatment in the second session. Ketamine and saline were administered during a 40min steady state infusion(27)and the sessions were at least 7 days apart. Participants were scanned 2h after the end of the infusion.

### Scales and questionnaires

The Psychotomimetic States Inventory (PSI) was used to assess the psychotomimetic symptoms that ketamine might produce (28) and completed at the end of each infusion. A greater PSI score indicates more drug-induced psychotomimetic experiences.

The Snaith-Hamilton Pleasure Scale (SHAPS) was used to assess anhedonia at the beginning of each scanning session as well as 2h after each infusion (29). Higher SHAPS scores indicate higher levels of anhedonia present.

### Image acquisition and Preprocessing

All scans were acquired using a GE MR750 3-Tesla scanner and a 16-channel head coil. Functional scans were obtained using T2* sensitive gradient-echo echo-planar imaging (EPI) (repetition time [TR]= 2000ms, echo time [TE]=30ms, flip angle= 75°, field of view [FoV]=214mm, slice thickness=3mm, number of slices=42). The initial four volumes of each timeseries were discarded to minimise steady-state effects on the signal amplitude. A total of 414 volumes were analysed for each timeseries acquired. A T1-weighted MPRAGE scan (FoV= 204mm, TR=7.3ms, TE=3ms, 256×256×156 matrix, slice thickness=1.2mm) was acquired on each session and was used for the reconstruction of a DARTEL template (30).

All structural and functional data were analysed using SPM-12. Pre-processing steps included realignment of the scans for each session as well as between sessions, co-registration to the MPRAGE image and normalization using the DARTEL flow fields. The normalised images were then smoothed using an 8mm FWHM kernel. During the first level modelling, the six motion parameters estimated during the realignment were used as regressors along with frame wise displacement (31). One participant was excluded from the analysis due to excessive movement.

### The MID task

The version of the task closely followed that described in Knutson et al., 2001 with a detailed description in supplemental materials. The task consists of 96 trials of different reward magnitudes (High win trials, Low win trials, Neutral trials), signalled by the initial cue. The cue image is followed by a variable delay after which a target appears on the screen and participants have to respond with a left button press. During the feedback phase, the outcome of the trial and the total amount won are presented to participants. For the anticipation phase of the task, three regressors were created corresponding to different reward magnitudes associated with the task cues: “High win anticipation”, “Low win anticipation” and “Neutral anticipation”. The feedback phase of the win and no-win trials of the task were modelled separately and four regressors were created: “High win feedback”, “Low win feedback”, “High no-win feedback” and “Low no-win feedback”. All the anticipation and feedback contrasts were examined separately for the ketamine and placebo session and compared between the two drug conditions.

### Regions of interest definition

The ROIs we selected comprised the amygdala, the ventral and dorsal striatum, the VTA and the insula. The bilateral ROI for the ventral striatum (NAc) was defined as described in Montgomery et al., (2006), based on previous work from Mawlawi et al., (2001). The amygdala, the dorsal striatum, the VTA and the insula were anatomically defined using the FSL Harvard-Oxford atlas (34). All ROIs were thresholded for grey matter with the minimal probability index set at 20% and binarized. The mean beta estimates from the first-level modelling were extracted for each ROI using MarsBar. The ROI values were extracted for each subject and for each contrast, for the ketamine and placebo sessions and were analysed in SPSS version 25.

### Ketamine’s metabolites

Blood samples were collected at the beginning of each study session, immediately after the drug infusion, and 2h after the end of the infusion. Ketamine, norketamine and the two isoforms of hydroxynorketamine ((2R, 6R)-HNK; (2S, 6S)-HNK) were measured in these samples. The values were used as a correlates with the ROI data to explore whether changes in brain activations induced by ketamine were related to the plasma exposure to ketamine and its main metabolites.

### Statistical analyses

The overall effect of treatment on each task contrast was examined using a mixed-effects model in SPSS. Each contrast was explored further by comparing the ROI activation between ketamine and placebo using a paired t-test and within each treatment session by using a one-sample t-test. Bonferroni correction for multiple comparisons was applied (p=0.008). In order to examine whether the ketamine metabolite levels, 2h post infusion, would predict the ROI activation under ketamine, we performed robust regressions. The placebo beta values were used as a covariate in this analysis to account for individual differences in brain activations and FDR correction was applied.

## Results

### Subjective Effects of ketamine

The established increase in psychotomimetic effects on ketamine were shown with the psychotomimetic states inventory (PSI) total score (Ketamine: mean=48.4, SD=±22.9, Placebo: Mean=15.1, SD=±10.6) and six subscales (see Figure 1). The immediate effects of ketamine were as expected, and the low placebo scores also aligned with expectations for this group of remitted depressed volunteers who did not experience any significant symptoms including anhedonia. This was also confirmed by the SHAPS, which as expected, indicated very low levels of anhedonia pre-infusion (pre-placebo mean score=22.7, SD=±5.6, pre-ketamine mean score =21.8, SD= ± 5.4, Wilcoxon signed test, Z=-0.811 p>.05) that remained unchanged after ketamine (2h post-ketamine mean score=21.9, SD=±5.3, Wilcoxon signed rank test, Z=-0.981 p>.05).

**Figure 1.**
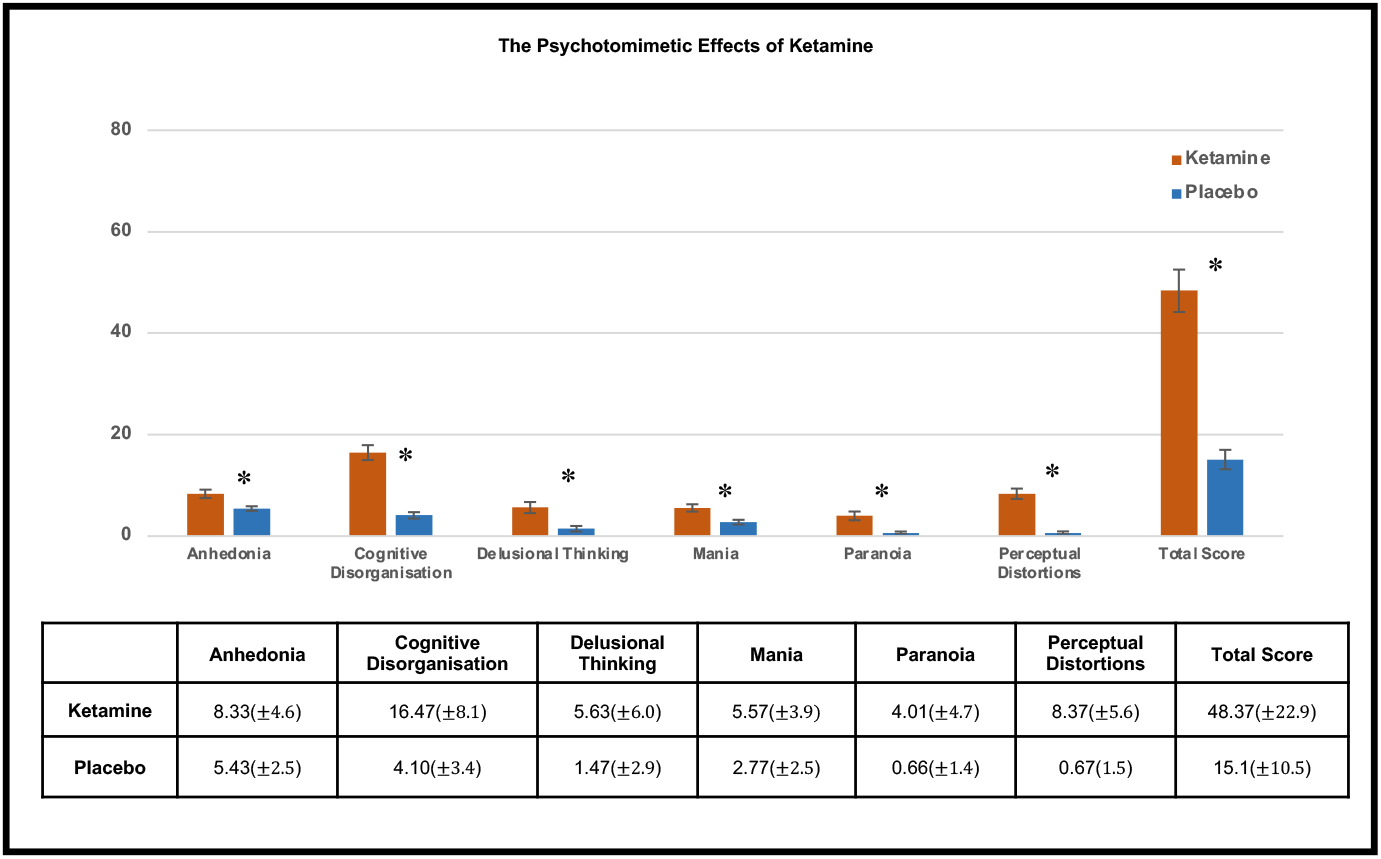
Ketamine administration produced robust psychotomimetic effects as measured by the PSI, right after the infusion. Significant increases were observed in the total score as well as the six PSI subscales for the ketamine session compared to the placebo session (paired t-test, p<.05).

**Figure 2.**
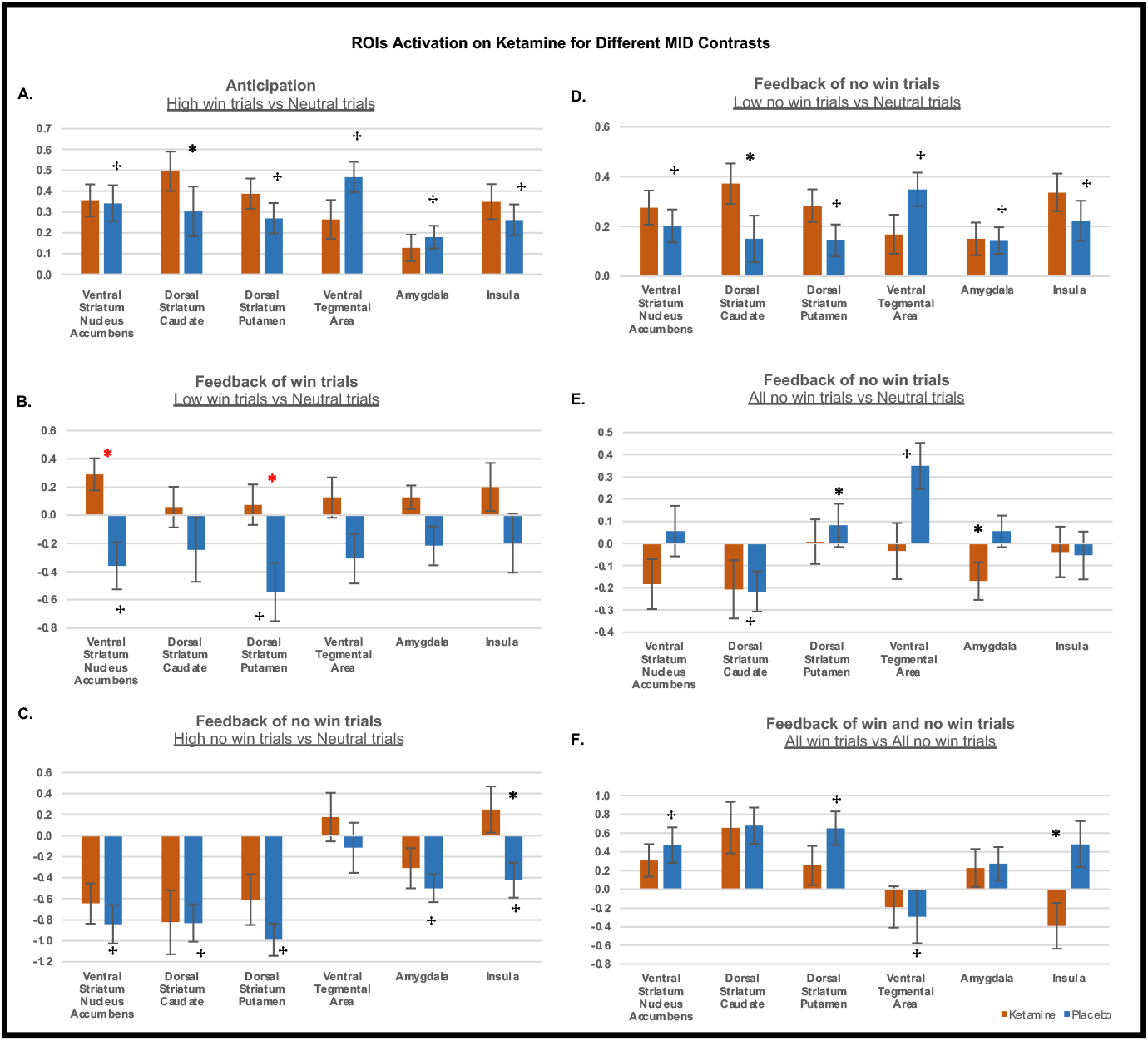
The activation of our pre-defined ROIs was examined for the anticipation (A) and feedback phase of the high and low win and no win trials (B-F). The beta values extracted for each contrast were compared between the ketamine and placebo sessions. All significant comparisons (paired t-test, p<.05) are indicated with an asterisk. When the feedback phase of the low win trials was contrasted to the feedback phase of neutral trials the ventral striatum/nucleus accumbens and the dorsal striatum presented with significant increases 2h post ketamine compared to placebo (B) and this result survived Bonferroni correction for multiple comparisons (p_CORR_ = 0.008), indicated with a red asterik. The ROIs that were significantly activated (p_FDR_CORR_<.05) for the same contrast in the placebo session alone are indicated with a cross. The task activations under placebo are presented in more detail in the Supplementary Material

**Figure 3.**
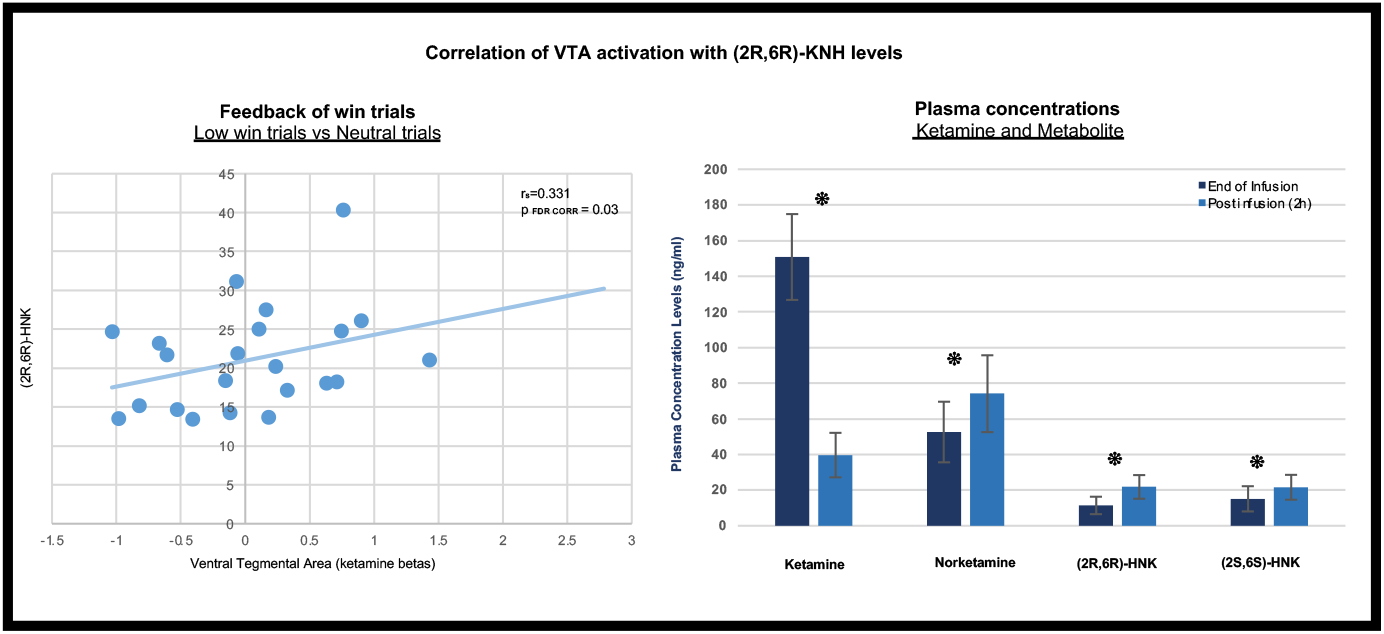
A.The levels of (2R,6R)-HNK, as measured 2h post-infusion, significantly correlated (r_s_= 0.33, p= 0.03) with the activation (beta values) of the Ventral Tegmental Area during the ketamine session and when the feedback phase of low win trials was contrasted to that of neutral trials. This finding remained significant (p = 0.033) when a robust regression was performed using the placebo beta values as a covariate to account for individual differences in brain activation during that contrast. B. The blood concentrations for ketamine and its main metabolites were measured at the end of the 40min infusion and 2h post infusion.

### The MID task

#### Task performance

The total amount of money won during the task did not significantly differ (Paired t-test, p>.05) between the ketamine and placebo sessions (Ketamine: 45.1, SD=±5.5, Placebo: 43.3, SD=±9.1). For reaction times there was a main effect of reward magnitude with faster responses for high win trials (F(2,36)=23.2, p<.0001) and no interaction with drug.

### Brain activations on placebo

The brain activations during the anticipation and feedback phases of the MID task, aligned with expectations based on previous studies (see Supplementary Figure 1).

### Ketamine’s effects on the MID task

For the whole brain analyses there were no differences between the ketamine and placebo sessions.

The a priori defined ROIs were examined for all the contrasts that were created for the MID task, and here we present the specific contrasts for which ROI activation significantly changed between the ketamine and placebo sessions. The statistical values for the main effects are provided in the text, and for the ROIs, in the figures and legends.

### Anticipation phase

A main effect of ketamine increasing activity was identified for the anticipation of all win trials compared to neutral trials across the predefined ROIs (F(1,36)=9.261, p=.003). No main drug effect was identified when the anticipation phases of high and low win trials compared to neutral trials were examined separately or compared to each other. Individual ROIs were not significant after multiple comparisons correction for the number of ROIs (Figure 1A).

### Feedback phase – win trials

A main effect of ketamine increasing activity was identified for the feedback phase of low win trials compared to neutral trials (F (1,36)=4.563, p<.001).

When the feedback phase of win trials was explored further, ketamine, compared to placebo, increased activations in the NAc and the putamen during the feedback phase of low win trials compared to neutral trials (Figure 1B). This effect survived correction for multiple comparisons.

### Feedback phase – no-win trials

A main effect of ketamine was observed when the feedback phase of all the no-win trials was contrasted to the neutral trials (F(1,36)=5.467, p<.001) and when the feedback phase of high no-win trials was compared to neutral trials (F(1,36)=5.859, p=0.016). For individual ROIs none of these effects survived correction for multiple comparisons (Figures 1C-E).

### Feedback phase-win trials vs no-win trials

A main effect of ketamine was identified when all the win trials were compared to the no-win trials (F(1,36)=5.036, p<.001), but no single ROI showed a significant change by itself after correction for multiple comparisons (Figure 1F)

### Association of ROIs activation with (2R,6R)-HNK levels

A positive correlation was identified, using robust regression, between the VTA activation, 2h post ketamine and the plasma levels of (2R, 6R)-HNK, 2h post the ketamine infusion (n=22, p_FDR_= 0.03). This correlation was identified when the feedback phase of low win trials was contrasted to that of neutral trials (Figure 4). A positive correlation was also identified for the activation of the caudate, 2h post ketamine and the plasma levels of (2R, 6R)-HNK when high no win trials were contrasted to neutral trials. This finding did not survive testing for multiple comparisons.

There were no relationships between ROI values and ketamine, norketamine and (2S, 6S)-HNK plasma levels for any of the task contrasts.

## Discussion

Ketamine, approximately 2h after its administration, modulated brain activity during the MID task, in areas that are important for reward processing. To our knowledge our study is the first to demonstrate that ketamine can produce detectable changes in the activation of brain areas that are important for reward processing and anhedonia 2h after infusion, without concurrent changes in depressive symptoms and the confounding effects of antidepressant treatment.

Previous studies have shown that ketamine, 24h after its administration normalises some of the connectivity changes observed in depression (35, 36) as well as reducing hyperactivation in the sgACC during a reward processing task (37). All these effects, at the time-point when they were observed, were accompanied by improvements in depressive symptoms and thus could either be attributed to the primary effects of the drug on neural processes that are affected in depression or could be the secondary effect of symptom changes that ketamine produces. In our cohort of remitted depressed volunteers, depressive symptoms and anhedonia were not present and did not change with ketamine suggesting that the drug can directly modulate reward-related neural processes (17, 18, 21) producing differential effects depending on the task contrast.

Ketamine increased the activation of the NAc, the putamen, the insula and the caudate when the feedback phase of win and no-win trials was compared to that of neutral trials (Figure 1B-1D). Recent meta-analyses have shown that striatal regions present with decreased activations during the anticipation and feedback phase of the MID task in patients with a mixture of mood disorders (2, 12). Moreover, striatal hypofunction persists during remission (15) and altered brain activations in those areas could also contribute to the blunted responses to positive feedback that characterises remitted depressed individuals (38). Remitted depressed and depressed individuals also demonstrate heightened neural responses to negative feedback (39) which has been related to anhedonia.

The fact that ketamine, during the feedback phase of the MID task, approximately 2h post-administration, altered the activation within the mesolimbic reward pathway provides a plausible mechanism by which ketamine could modulate abnormal responses to positive and negative feedback. Several of the brain areas where ketamine-induced alterations were observed in our study are also target areas for antidepressant treatments with different pharmacology (40) and changes in their activation and connectivity predicts treatment response (41, 42). Taken together these findings indicate that the effects observed in our study, 2h post ketamine, could be relevant to symptoms’ improvement in depression. However, in order to fully understand the consequence of these changes in the modulation of specific symptoms such as anhedonia and guilt (39), studies in actively depressed patients will be needed.

In our study, we found preliminary evidence to link the changes in brain activity with the levels of an active metabolite of ketamine, (2R, 6R)-HNK. The increases in brain activity in the VTA during the feedback phase of low win trials positively correlated with the levels of (2R, 6R)-HNK. It has been suggested that direct activation of AMPA receptors by (2R, 6R)-HNK triggers the plasticity-related pathways, mediating ketamine’s antidepressant action (23). Brain areas of the mesolimbic pathway receive dense glutamatergic input and glutamate receptors of this pathway are crucial for synaptic plasticity (43). While there is no direct evidence of increased plasticity after ketamine in patients, PET studies support this conclusion through increased glucose metabolism, which correlates with improvements in depression symptoms and anhedonia in the VS, dACC and putamen 2h post-infusion (17, 18). Taken together with studies of Lally et al(17, 18)., our findings demonstrate the potential value of concurrent measurement of brain metabolism, functional modulation of brain activity, symptom changes and metabolites levels in building a model of the effects of ketamine in improving specific symptoms.

This study has a number of limitations. First, the absence of a healthy volunteer group does not allow the direct characterisation of impairments in reward processing in our remitted group and thus establish whether the effect of ketamine is towards a normalization of these changes. Second, most of the ketamine associated changes have been identified during the feedback phase of the MID task highlighting the role of positive and negative outcomes for reward processing and anhedonia. The strength of the MID task design is in the reward anticipation phase with fewer trials contributing to the feedback contrasts, thus future studies using a reward task designed to focus on outcomes will help in replicating the feedback effects, as well the potential relationships with anticipation effects. While it remains possible that the effects during feedback are a consequence of the drug effects during anticipation, this is unlikely as both increases and decreases in activity were observed during feedback on ketamine versus placebo. These differential effects also do not fit with an interpretation of the drug effect being understood as a change in neurovascular coupling.

In summary, this study demonstrates that ketamine, 2h post administration, could produce detectable changes in brain areas that are part of the mesolimbic pathway involved in reward processing. These changes were not secondary to symptom changes in our cohort of remitted depressed volunteers. During the feedback phase of low win and high no-win trials, changes in brain activity correlate with the levels of (2R, 6R)-HNK. These findings support a model whereby ketamine improves reward processing deficits via enhanced anticipation of reward and modulation of responses to negative feedback, and also highlight the importance of the drug metabolite levels in understanding ketamine’s antidepressant and anti-anhedonic actions. Future studies examining the role of ketamine’s metabolites during reward processing task in depression would contribute to our understanding of ketamine’s antidepressant action.

Supplementary Information is available on MP’s website

## Data Availability

All the data referred to this manuscript are available upon contacting the corresponding author

## Acknowledgements

This work was supported with founding from Johnson and Johnson via a research grant awarded to University College London and King’s College London. This paper represents independent research part-supported by the National Institute for Health Research (NIHR) Biomedical Research Centre at South London and Maudsley NHS Foundation Trust and King’s College London. The views expressed are those of the author(s) and not necessarily those of the NHS, the NIHR or the Department of Health and Social Care. The authors of this paper would like to thank the Centre for Neuroimaging Sciences’ research stuff for all their hard work and support during throughout this research project.

## Conflict of Interest

Dr Curran’s research is supported by the UK Medical Research Council and NIHR; she has consulted for Jannsen on esketamine. Dr Mehta has received grant funding from J&J, Lundbeck and Takeda and has acted as a consultant for Lundbeck and Takeda. Dr Furey is an employee of Janssen Research and Development

## Notes

### Competing Interest Statement

The authors have declared no competing interest.

### Clinical Trial

NCT04656886

### Funding Statement

This work was supported by a research grant awarded by Johnson and Johnson awarded to UCL and King's College London

### Author Declarations

This research has been approved by the PNM Research Ethics Subcommittee of King's College London: KCL/REC 0650

